# *APOL1* genotype and patient outcomes in US and South African transplant recipients with HIV who received kidneys from donors with HIV

**DOI:** 10.1101/2025.04.15.25325856

**Authors:** Robert Freercks, Moreno Rodrigues, Kathryn Manning, Jurgen Heymann, Jeffrey B. Kopp, Savania Nagiah, Meenakshi Rana, Sander Florman, Rachel Friedman-Moraco, Peter Stock, Alexander Gilbert, Shikha Mehta, Valentina Stosor, Marcus R Pereira, Michele I Morris, Jonathan Hand, Ghady Haidar, Maricar Malinis, Carlos A. Q. Santos, Joanna Schaenman, Emily A. Blumberg, David Wojciechowski, Jonah Odim, Allan Massie, Miruthula Tamil Selvan, Serena Bagnasco, Dorry Segev, Aaron AR Tobian, Elmi Muller, Christine M Durand, Andrew D Redd

## Abstract

**Importance:** Lower kidney allograft survival has been demonstrated in kidney transplant recipients (KTR) without HIV whose donors have two apolipoprotein L1 (*APOL1*) renal risk variants (RRV). The effects of *APOL1* RRV on kidney transplant outcomes in people with HIV (PWH) have not been fully assessed.

**Objective:** To determine whether *APOL1* renal risk variants (G1/G2) in donors or recipients are associated with outcomes of kidney transplantation in people with HIV (PWH)?

**Design:** Comparative analysis of kidney allograft outcomes in two of the largest longitudinal clinical studies examining transplantation outcomes in PWH.

**Setting and participants:** Two cohorts of HIV-positive KTR (R+) and their respective HIV-negative (D-) or HIV-positive (D+) kidney donors from the South African (SA) HIV+ to HIV+ transplantation clinical study and the United States of America (US) HOPE in Action Kidney transplantation clinical trial. All patients with genomic DNA available for *APOL1* genotyping were included. *APOL1* Genotype was determined using a probe-based assay.

**Main outcomes measured:** Time to first rejection, HIV-associated nephropathy, graft failure or death were compared by both donor and recipient *APOL1* RRV status.

**Results:** Genomic DNA was available for 21 donors with HIV and 38 HIV D+/R+ recipients in the SA cohort, and 57 donors (40 D+ and 17 D-) and 119 recipients (49 HIV D+/R+ and 70 D-/R+) in the US cohort. Recipient outcomes were not associated with recipient *APOL1* genotype. However, recipients whose donor carried one versus zero *APOL1* RRV were significantly more likely to experience a negative composite outcome (p<0.02 for both cohorts independently), which led to an adjusted hazard ratio of a poor composite outcome of 2.9 (95% CI 1.1–7.4) and 10.1 (95% CI 2.4–42.7) in the SA and US cohorts, respectively.

**Conclusions and relevance:** In two independent studies, the presence of one *APOL1* RRV in a donor kidney led to significantly worse post-transplant outcomes while recipient *APOL1* genotype was not associated with outcomes. Further research into the interaction between the allograft environment and donor *APOL1* genotype in PWH is required.

**KEY POINTS:** 

**Question:** Do *APOL1* renal risk variants (G1/G2) influence the outcomes of kidney transplantation in people with HIV (PWH)?

**Findings:** In two of the largest cohorts of PWH who are also kidney transplant recipients, the presence of even one donor *APOL1* renal risk variant was associated with an adjusted hazard ratio of a poor composite outcome of 10.1 (95%CI=2.4-42.7) and 2.9 (95%CI=1.1-7.4) in the US and SA cohorts, respectively. Recipient *APOL1* genotype was not associated with graft outcomes.

**Meaning:** This may have implications for allocation of allograft kidneys in PWH, as well as informing the need for therapies targeting *APOL1* gene expression in kidney transplant recipients.

## Introduction

The outcomes of kidney transplantation in people with HIV (PWH) with controlled HIV replication are equivalent to those of people without HIV ^1^, and for recipients with HIV regardless of donor HIV status ^2^. Accordingly, kidney transplantation from donors with HIV to recipients with HIV (HIV D+/R+) is now a standard care option in the United States of America (US)^3^. While this has already reduced organ waiting times for PWH ^4^, as well as reducing waitlists overall, there remains a need to further improve long-term graft function and survival.

The discovery of apolipoprotein L1 gene (*APOL1*) risk alleles as contributors to genetic risk for kidney disease in those of recent African ancestry, has fundamentally altered our understanding of the contribution of continental ancestry to progressive chronic kidney disease (CKD) ^5^. These *APOL1* renal risk variants (RRV; labelled G1: S342G and I384M; and G2: del N388 and Y389) afford protection against trypanosomiasis seen in Central and West Africa, but are also associated with progressive CKD and significantly contribute to genetic risk of CKD, particularly in PWH, in whom penetrance is the highest ^6^. These RRV are largely absent in Americans of European descent and are significantly more common in West Africans ^7^ and African Americans ^8^. In US population-based studies, approximately 37% of African Americans carry one RRV while 13% carry two ^9^. The G1 and G2 RRV are also present in South African populations, with population frequencies of 7.3% - 10.0% and 14.9% - 23.7% respectively ^10^.

Locally-synthesised APOL1 protein variants mediate kidney toxicity through various mechanisms, including induction of mitochondrial and other intracellular toxicities in kidney podocytes, in part through a pore-forming mechanism, manifested especially in individuals with two *APOL1* RRV in any combination ^11^. The expression of *APOL1* is upregulated by interferon gamma ^12^, whose expression is also increased in HIV and may represent the link between *APOL1* RRV carriage and harmful expression leading to CKD ^13^.

Donor *APOL1* RRV are also associated with shorter renal allograft survival in KTR without HIV, and this helps to explain the shorter allograft survival seen in African American KTR ^14^. The utility of pre-implantation *APOL1* genotyping is currently uncertain, and further studies are required to better evaluate such an approach ^15^. The presence of donor or recipient *APOL1* RRV in HIV-positive deceased donor kidneys may influence renal allograft survival in PWH who receive kidney transplants.

To date, there are a lack of data on the influence of *APOL1* RRV and outcomes of kidney transplantation in PWH, where the additional presence of HIV may significantly influence *APOL1* expression. A better understanding of the relative importance of donor versus recipient *APOL1* genotype may lead to improved overall outcomes. We aimed to determine this in two independent cohorts of kidney transplantation in PWH, one in South Africa (SA) and one in the US.

## Methods

### South African study cohort

The SA population in this study has been described previously ^16,17^. Briefly, SA KTR with HIV receiving kidneys from deceased donors with HIV (HIV D+/R+), and their respective donors, had peripheral blood mononuclear cells (PBMC) collected, processed and frozen at −112°F prior to transplantation and at several follow-up time points, which were subsequently used for *APOL1* genotyping. When available recipient pre-transplant samples were used for genotyping. All recipients and donors that had an available PBMC sample were tested for this study. The groups analysed here were not significantly different than the full cohort (Table S1 & S2). All participants provided informed consent, and the study was approved by the University of Cape Town Human Research Ethics Committee (UCT HREC 927/2014).^1^

### United States study cohort

The US population in this study was taken from the HOPE in Action Kidney transplantation clinical trial, which examined the safety and efficacy of HIV D+/R+ transplantation vs donation from donors without HIV (HIV D-/R+) ^2^. This population has been described in detail previously ^2^. Briefly, between April 2018 and September 2022, 198 individuals with HIV underwent kidney transplantation under the multi-center observational HOPE in Action study (ClinicalTrials.gov: NCT03500315). There were 99 transplants in each trial arm (HIV D+/R+ and D-/R+). Similar to the SA cohort, recipients and donors from both HIV D+/R+ and D-/R+, had peripheral blood mononuclear cells (PBMC) collected, processed and frozen at −112°F prior to transplantation, and at several follow-up time points. The pre-transplant PBMC aliquots were subsequently used for APOL1 genotyping. All recipients and donors with available PBMC for genomic testing were included for this analysis. The groups analysed here were not significantly different than the full cohort (Table S3 & S4). All participants provided consent, and the study was approved centrally by the Johns Hopkins University School of Medicine Institutional Review Board (IRB00141138) and by each local transplant center.

### APOL1 genotyping

Genomic DNA was extracted from PBMC obtained from deceased kidney donors and recipients in both cohorts using the DNeasy Blood and Tissue kit (Qiagen, Hilden, Germany) following the manufacturer’s protocol. Samples from both cohorts were analysed at the same US National Institutes of Health laboratory using custom TaqMan probe-based assays specific for *APOL1* G1 SNPs (rs73885319 and rs60910145) and the G2 deletion (rs717185313) (ThermoFisher Scientific, Waltham MA, US)^18^. Non-risk *APOL1* RRV are designated as G0, G1, and G2 genotype calls were visually inspected for quality control. All samples showed consistent allele calls with 100% concordance across triplicate samples and only allowed genotypes were observed.

### Statistical analyses

Variables were compared by *APOL1* RRV status using the Mann-Whitney (Wilcoxon rank-sum test) for numerical variables (age, median CD4 count) and Fisher’s exact test for categorical variables (sex, self-reported race, outcomes). Kaplan-Meier survival curves were used to illustrate composite survival by *APOL1* RRV status and compared using log-rank test. The Cox proportional hazards model was used to generate hazard ratios and its 95% confidence interval (CI) to show the magnitude of effect for the presence of one *APOL1* RRV compared to none. Model was adjusted for age, hypertension and diabetes melitus and proportional hazard assumption was evaluated using Schoenfeld residual plots. Each cohort was examined individually, as well as in a combined analysis.

## Results

Genomic DNA was available for 21 donors and 43 recipients from SA (HIV D+/R+), 38 of whom had matched donor genotyping available (**Table 1**). In the US cohort, genomic DNA was available for 119 recipients, all of whom were PWH (**Table 1**). Of these, 72 had donor genotyping available, of whom 49 were HIV D+/R+ (**Figure S1**). Overall, recipients in both cohorts were older than donors, and while evenly split between male (51%) and female (49%) in the SA cohort, recipients were predominantly male in the US cohort (86%). Most recipients in both cohorts were Black African or African American, while donors were majority male and had significantly greater racial diversity. When comparing the US and SA cohorts, US donors also had a higher CD4 count, were more likely to be white race and less likely to carry *APOL1* RRV; this was similar when restricting the comparison to US donors with HIV (**Table S5-S7**). Donors from both cohorts had relatively low rates of dual *APOL1* RRV carriage with only one donor in the US cohort and no donors in the SA cohort carrying two *APOL1* RRV (Table 1). Conversely, recipients in both cohorts had significantly higher rates of *APOL1* RRV carriage, with 39% and 42% carrying two RRV in the SA and US cohorts respectively. HIV-associated nephropathy (HIVAN) was the original cause of native kidney failure in 15 of the 17 (88.2%) KTR with two *APOL1* RRV in the SA cohort and in 11 of 51 with two *APOL1* RRV (21%) in the US cohort.

**Table 1:** HIV-positive kidney transplant recipients and donor characteristics in South African and United States HOPE cohorts.

|  | South African cohort |  |  | United States HOPE cohort |  |  |
| --- | --- | --- | --- | --- | --- | --- |
|  | HIV<sup>+</sup><br>donors<br>N=21 | HIV<sup>+</sup><br>recipients<br>N=43 | P value | HIV<sup>+</sup>/HIV<sup>-</sup><br>donors<br>N=99 | HIV<sup>+</sup><br>recipients<br>N=119 | P value |
| **Demographic variables** |  |  |  |  |  |  |
| **Age (years)** | 31 (26-40) | 37 (33-48) | 0.006 | 36 (28-47) | 55 (46-63) | <0.001 |
| **Race** |  |  | 0.004 |  |  | <0.001 |
| Black or African American | 12 (57) | 39 (91) |  | 30 (30) | 84 (71) |  |
| Other | 7 (33) | 4 (9) |  | 21 (21) | 10 (8) |  |
| White | 2 (10) | 0 (0) |  | 48 (49) | 25 (21) |  |
| **Sex, male** | 16 (76) | 22 (51) | 0.064 | 69 (70) | 102 (86) | 0.007 |
| **HIV positive** | 21 (100) | 43 (100) |  | 46 (46) | 119 (100) |  |
| **CD4 (cells/mm3)** | 257 (33-805) | 447 (303-578) | 0.140 | 406 (284-810) | 526 (384-686) | 0.256 |
| **APOL1 risk alleles** |  |  |  |  |  |  |
| **No. of risk alleles** |  |  | <0.001 |  |  | <0.001 |
| 0 | 14 (77) | 21 (49) |  | 50 (88) | 44 (37) |  |
| 1 | 7 (33) | 5 (12) |  | 6 (11) | 24 (20) |  |
| 2 | 0 (0) | 17 (39) |  | 1 (1) | 51 (43) |  |
| **Haplotypes<sup>a</sup>** |  |  |  |  |  |  |
| G0/G0 | 14 (67) | 21 (49) |  | 50 (88) | 44 (37) |  |
| G0/G1 | 4 (19) | 4 (9) |  | 3 (5) | 8 (7) |  |
| G0/G2 | 3 (14) | 1 (2) |  | 3 (5) | 16 (13) |  |
| G1/G1 | 0 (0) | 7 (16) |  | 0 (0) | 28 (23) |  |
| G1/G2 | 0 (0) | 10 (23) |  | 0 (0) | 14 (12) |  |
| G2/G2 | 0 (0) | 0 (0) |  | 1 (2) | 9 (7) |  |

Data is presented as median (IQR) (continuous variables) or N (%) (categorical variables)
Abbreviations: *APOL1*, Apolipoprotein L1; <sup>a</sup>G0, Wild type genotype; G1, genotype S342G and I384M; G2, genotype del N388 and Y389

The median follow-up time after transplantation to first outcome was 3.58 (0.86-6.99) years and 1.60 (1.4-2.4) years in the SA and US cohorts respectively. Overall, five recipients (13.2%) in the SA cohort and 14 recipients (11.8%) in the US cohort died during the study periods (**Table 2**). The most common cause of death in both cohorts was infection, primarily due to COVID-19 pneumonia, which accounted for three (60%) and four (28.6%) of the deaths in the SA and US cohorts, respectively.

**Table 2:**
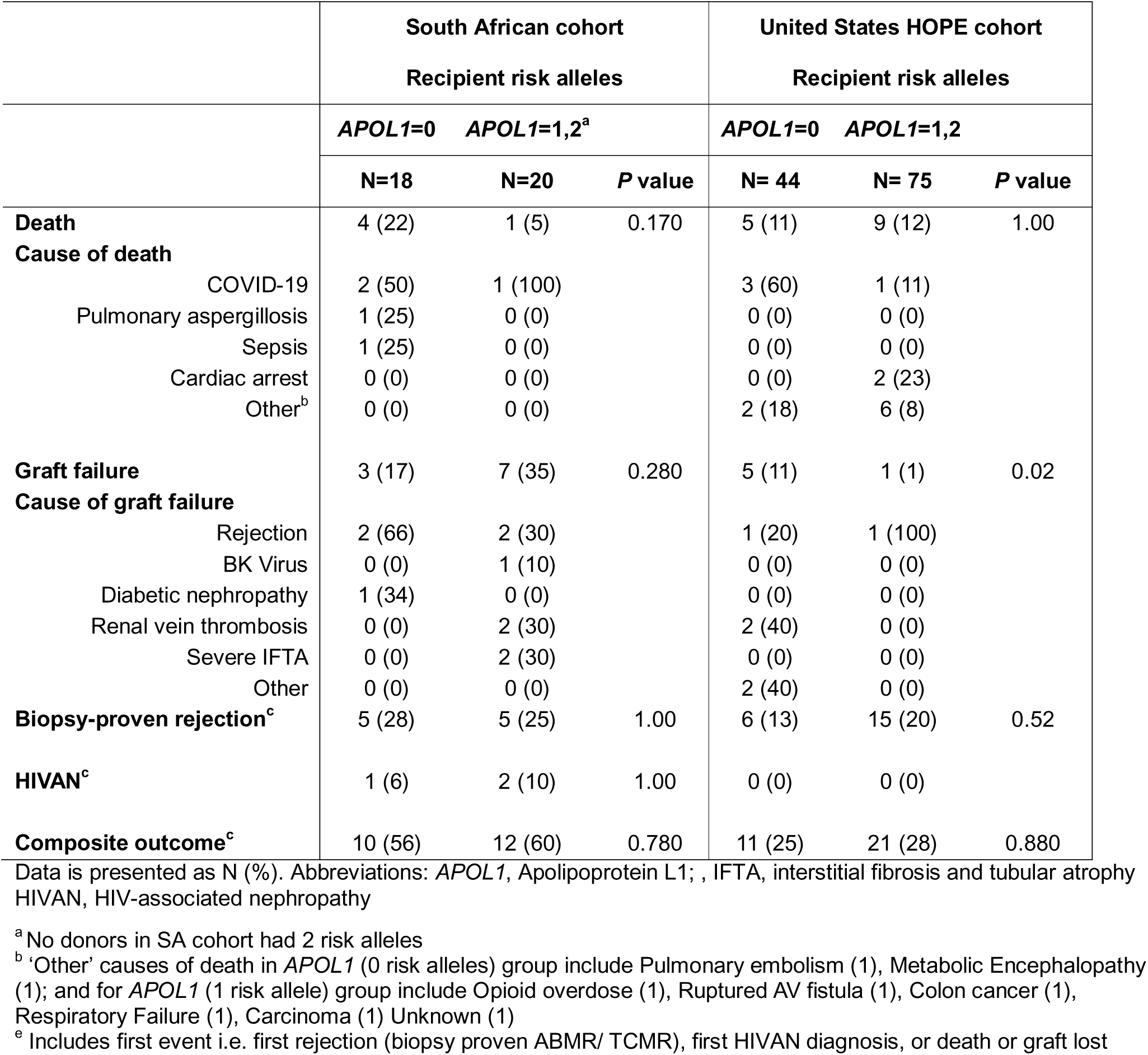
Recipient outcomes by *APOL1* recipient risk allele status in South African and United States HOPE cohorts.

The presence of one *APOL1* RRV in the donor was associated with a significantly increased likelihood of biopsy-proven kidney rejection (SA p=0.005 and US p=0.030) and death (US cohort only, p=0.001) for the recipient (**Table 3**). The composite of a first event (HIVAN, biopsy-proven rejection, graft failure or death) was observed in 11 of 12 SA recipients (92%) whose donor carried one *APOL1* RRV compared to 11 of 26 (42%) in those whose donor had zero *APOL1* RRV (p = 0.004; **Table 3**). Similarly, a poor composite outcome was noted in five of the seven US recipients (71%) whose donor carried one *APOL1* RRV compared to four of 42 (9%) whose donor had zero *APOL1* RRV (p = 0.001; **Table 3**). In the US cohort, there were no donors without HIV with *APOL1* RRV, so we could not determine the impact of HIV status on this association. Cox regression analysis showed a significantly increased risk of a poor composite outcome in KTR whose donors carried one versus zero APOL1 RRV, with hazard ratios of 8.36 (95% CI = 2.2–31.4) for the US cohort and 2.9 (95% CI = 1.2– 6.8) for the SA cohort. After adjusting for recipient age, diabetes and hypertension, the adjusted hazard ratio (aHR) of a poor composite outcome in KTR whose donor carried one versus zero *APOL1* RRV was 10.1 (95%CI=2.4-42.7) and 2.9 (95%CI=1.1-7.4) in the US and SA cohorts, respectively.

**Table 3:**
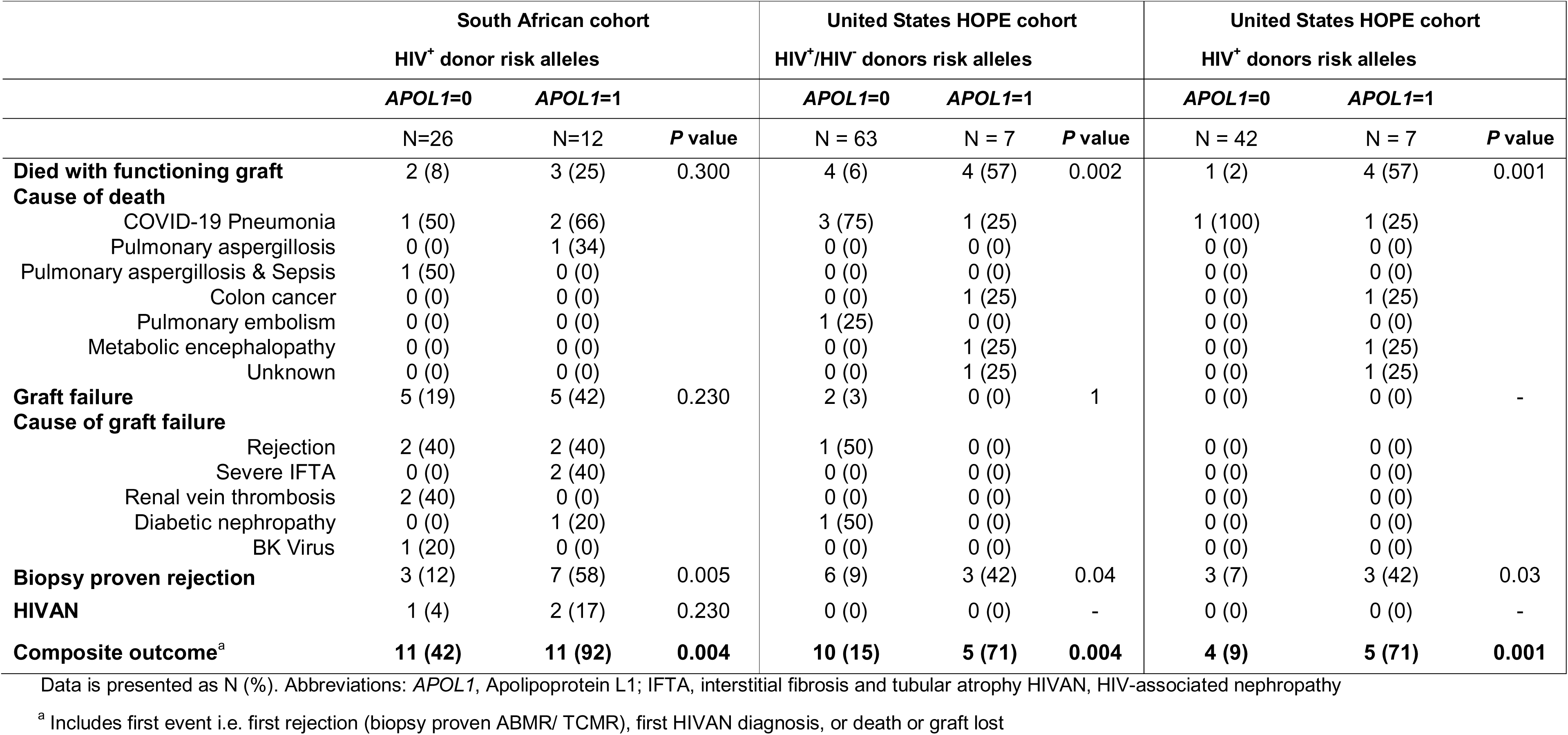
Recipient outcomes by donor *APOL1* risk allele status in South African and United States HOPE cohorts.

Kaplan-Meier estimations for a composite first event in KTR by donor *APOL* RRV status also demonstrated increased rates of a composite outcome of HIVAN, biopsy-proven rejection, graft failure or death in KTR of donor kidneys with one *APOL1* RRV in both the US and SA cohorts (p<0.02, **Figure 1**), as well as when the cohorts were combined for analysis (p<0.001; **Figure S2**). Similar findings were observed in the US cohort when including KTR who received kidneys from donors with one or two risk *APOL1* RRV (p=0.05), as well as when limited to donors with HIV (p=0.01; **Figure S3 and S4**). No recipient in the SA cohort received a kidney from a donor with two *APOL1* RRV, preventing us from evaluating this outcome. On sub-analysis of data combining both cohorts (**Figure S5A-D**), inferior graft outcomes were observed in KTR of donors with 1 *APOL1* RRV, driven primarily by increased rates of deaths (p<0.001) and rejection (p<0.001), but not for graft loss (p=0.76) or onset of HIVAN in the graft (p=0.101; **Figure S5A-D**).

**Figure 1:**
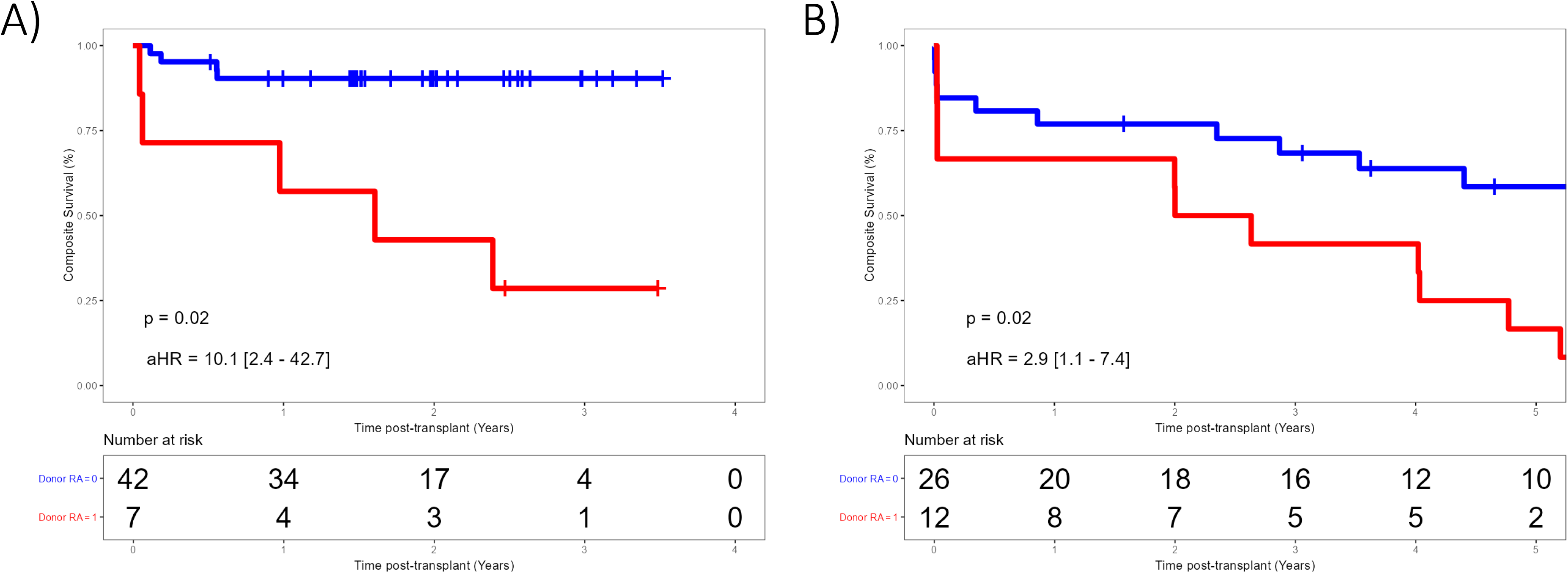
Kaplan-Meier estimations for a time to first outcome of a composite endpoint consisting of rejection (biopsy proven ABMR/ TCMR), HIVAN, graft lost or death) in kidney recipients by donor *APOL1* risk variant status in kidney recipients living with HIV in the US HOPE in Action (A) and the South African HIV D+/R+ (B) cohorts.

Conversely, recipient outcomes were not associated with recipient *APOL1* genotype (carriage of zero versus one or two *APOL1* RRV) and a composite of poor outcomes (first event of HIVAN, biopsy-proven rejection, graft failure or death) was similar in both groups (SA cohort: 10 of 18 [56%] versus 12 of 20 [60%], p=0.780; US cohort: 11 of 44 [25%] versus 21 of 75 [28%], p=0.880, **Table 2**).

## Discussion

In this study of PWH enrolled in two independent KTR cohorts in SA and the US, the presence of a single donor *APOL1* RRV was associated with highly similar findings in both cohorts of significantly worse outcomes in the renal allograft recipients compared to recipients receiving kidneys from donors with zero *APOL1* RRV. Although dual *APOL1* RRV carriage in KTR was common in both cohorts, the presence of either one or two *APOL1* RRV in KTR was not associated with worse kidney allograft outcomes compared to KTR with zero *APOL1* RRV. The two cohorts were similar in their baseline demographic characteristics, although recipients in the US tended to be older and to have greater racial diversity. US donors were also less likely to carry *APOL1* RRV, in keeping with a lower proportion of participants with recent African ancestry.

In the general population, APOL1-mediated kidney disease (AMKD) follows a largely recessive inheritance pattern. Similarly, studies investigating the influence of *APOL1* on transplant outcomes have largely focused on the effect of two *APOL1* RRV in kidney donors ^19^. However, a recent large prospective cohort study of CKD from West Africa demonstrated a clear gene dose effect, with any single *APOL1* RRV (G1 or G2) being associated with an 18% higher odds of CKD and a 61% higher odds of having focal segmental glomerulosclerosis ^7^. This is in keeping with other smaller studies that have also demonstrated a small increase in risk for CKD in *APOL1* RRV heterozygotes ^20^. This may be especially important in the setting of kidney transplantation, where the graft kidney is exposed to additive environmental insults including traumatic injury leading to brain death, cold ischemic time, reduced functional kidney mass, and exposure to nephrotoxic medications. Recently, stabilization of hypoxia-inducible transcription factors by hypoxia has been shown to activate transcription of *APOL1* in podocytes and tubular cells ^21^. Furthermore, within the context of HIV infection, the recipient environment may be especially primed for injurious *APOL1* expression, with consequent AMKD. *APOL1* is an interferon-stimulated gene, and even with complete viral suppression in HIV, as in the present study, persistent immune dysregulation and enhanced interferon production is evident, which we speculate may upregulate expression of even one RRV and result in AMKD in this setting ^22^.

The presence of one *APOL1* RRV was associated with increased risk for allograft rejection in both cohorts, and to our knowledge this association has not been reported previously. Given the relatively small size of this study, these findings should be interpreted with caution. In a large cohort examining the presence of focal and segmental collapsing glomerular lesions typical of AMKD in KTR who were carriers of two *APOL1* RRV, concomitant rejection was seen in 61% of patients ^23^. In the US cohort, there was also an association between COVID-19 related mortality and *APOL1* RRV. This agrees with the findings by Hung et al. who recently reported the association of dual *APOL1* RRV carriage with acute kidney injury and death in a large cohort of African American veterans hospitalized with COVID-19 ^24^.

The present findings are in keeping with other studies that have shown an association between graft failure and *APOL1* RRV carriage by donors rather than by recipients. In one study of 136 African American deceased donor kidneys and their respective recipients, the presence of two *APOL1* RRV in donors was more important in determining outcomes than cold ischemic time or HLA mismatch. The same study also showed that worse allograft outcomes with African American donor kidneys could be solely attributed to donor *APOL1* genotype rather than to race ^25^.

Similar to the present study, another small retrospective study of 119 African American kidney transplant recipients reported that recipient carriage of two *APOL1* RRV was not associated with inferior transplant outcomes ^26^. However, one study did demonstrate inferior graft outcomes at one year in recipients with two *APOL1* RRV, although donor genotype was not reported in the study and outcomes were similar at three years ^27^. Zhang et al also recently described an increased risk of death-censored allograft loss and rejection in recipients with one or two *APOL1* RRV’s and this was independent of donor genotype ^28^. Interestingly, the same study also showed that *APOL1* RRV may alter immune cell function, in particular T cell and NK cell function, which could have an impact on rejection. Uncertainty remains regarding the relative importance of donor versus recipient *APOL1* genotypes and further research is currently underway to explore this question ^29^.

Our findings have important implications for transplantation in PWH, but will require confirmation in a larger study. In this regard, the National Institutes of Health-sponsored APOL1 Long-term Kidney Transplantation Outcomes Network (APOLLO) is prospectively assessing kidney allograft survival from donors with recent African ancestry based on donor and recipient *APOL1* genotypes and includes PWH ^29^. Strategies to prevent AMKD in the setting of HIV have not been tested, but could include selection of antiretroviral therapy shown to reduce interferon expression ^30^. Additionally, phase 2/3 studies of inaxaplin, a small molecule inhibitor of AMKD are currently underway and may be a promising adjunct in KTR whose donors carry *APOL1* RRV ^31^.

While the role of screening donors and recipients for *APOL1* RRV is currently unclear and complicated by the need to consider *APOL1* haplotype, as well as any potential unintended consequences such as implications for health insurance or stigmatization, the British Transplantation Society has recently recommended offering *APOL1* genotyping to all potential living donors. As therapies directed against AMKD become available, the role of testing may become clearer.

The present study has limitations. First, the study populations in each cohort were relatively small, but the identical findings in two different cohorts, as well as the combined data, suggest that the findings are robust. We also only had one donor with two *APOL1* RRV, and no donors without HIV with one or two *APOL1* RRV, limiting our ability to assess any increased risk afforded by these factors. Second, we were unable to test for additional *APOL1* haplotypes, and it is possible that such data may alter our understanding of variant risk and the role of one versus two *APOL1* RRV. One major strength of the present study was the ability to compare and combine the two largest independent cohorts of HIV D+/R+ kidney transplantation on different continents over a similar study period.

## Conclusion

In summary, we have shown that kidney allograft outcomes are worse in PWH who receive kidneys from deceased donors with one *APOL1* RRV, while recipient *APOL1* genotype did not influence graft outcomes. The mechanisms for the former remain unclear, although it is possible that toxic expression of *APOL1* is enhanced in KTR who also have HIV and by the transplant process itself, including concomitant medications. These findings should be confirmed in a larger cohort and further research into the interplay between the allograft environment and donor *APOL1* genotype in PWH is required.

## Data Availability

All data produced in the present study are available upon reasonable request to the authors.

## Acknowledgements

The authors would first like to thank the patients, donors and their families, and clinical teams who made this work possible. We would also like to thank the full HOPE in Action team (see list in supplemental data).

## Author contributions

RF, ADR – Wrote first draft of paper; JH, JBK, MTS – performed and oversaw laboratory work; RF, ADR, EM, CMD, AART - planned the laboratory study; CMD, DS, AART, ADR, JO – planned and oversaw the US transplant study; EM, ADR, KM – planned and oversaw the SA transplant study; MR, KM – analyzed primary data; MR, SF, RFM, PS, AG, SM, VS, MRP, MM, JH, GH, MM, CS, JS, EB, DS, CMD, AART – US cohort clinical team; SB – reviewed US pathology; RF, AR, CD and EM interpreted the data; and all authors reviewed and approved the final manuscript.

## Supplemental figure legends

**Figure S1:** Flow of US HOPE cohort. * indicates one donor with 2 APOL1 RRV removed for purposes of comparison.

**Figure S2:** Kaplan-Meier estimation for a composite first event in KTR by donor *APOL* RRV status in combined US and SA cohorts

**Figure S3:** Kaplan-Meier estimation for a composite first event in KTR by donor *APOL* RRV status in US HOPE cohort including all donors (HIV+ and HIV-)

**Figure S4:** Kaplan-Meier estimation for a composite first event in KTR by donor *APOL* RRV status in US HOPE cohort limited to HIV+ donors

**Figure S5:** Kaplan-Meier estimation for first event of death (A), biopsy-proven rejection (B), graft loss (C), and HIVAN (D) in KTR by donor *APOL* RRV status in combined US and SA cohorts

## Data sharing statement

Data will be made available upon request with approval and appropriate data use agreements. Please send requests for data from the US study to, and from the South African study to.

## Additional acknowledgements for HOPE in Action team

University of Alabama at Birmingham: Katherine Basinger, RN, CCRP; Darnell Mompoint Williams; DNP

University of Arkansas for Medical Sciences: Emmanouil Giorgakis, MD

University of California, San Diego: Layla Myers

University of California, San Francisco: Ada Chao, Joanne Kwon

University of California, Los Angeles: Adreanne Rivera

Yale University School of Medicine: Ricarda Tomlin, BS, CCRP

MedStar Georgetown Transplant Institute: Rochell Yacat

Miami Transplant Institute: Shweta Anjan, MD, Isabel Vital, Carlos Munoz, Lissett Moni

Emory University: Jeryl Huckaby, MSCRA, CCRC; William Kitchens, MD

Northwestern University: Michelle Callegari, Zachary C Dietch, MD.

Rush University Medical Center: Rebecca Lai

University of Illinois at Chicago: Kelly Bruno

Indiana University: Mary Balmes-Fenwick

Ochsner Clinic Foundation: Angela R. Smith, MBA

Massachusetts General Hospital: Kerry Crisalli, RN

Johns Hopkins University: Jamie Wiles, RN

University of Maryland, Institute of Human Virology: John Baddley, MD; Lisa Anderson

Columbia University Medical Center: Dominique Piquant

Icahn School of Medicine at Mount Sinai: Brandy Haydel, CCRC

Weill Cornell Medical College: Britta Witting

NYU Langone Transplant Institute: Rebecca Dieter, PharmD

University of Cincinnati: Senu Apewokin, MD, FACP; Jenny Baer, RN

University of Pittsburgh Medical Center: Ellie Morell

University of Pennsylvania: Maryann Najdzinowicz, RN

Methodist Health System Clinical Research Institute: Jose A. Castillo-Lugo, MD; Karen Castro

University of Texas Southwestern Medical Center: Ricardo M. La Hoz, MD; Jarrett Hubbard

