## Supplemental Tables S1 - S7 for "*APOL1* genotype and patient outcomes in US and South African transplant recipients with HIV who received kidneys from donors with HIV"

**Table S1:** Comparison between SA kidney recipients that are included or not included in study

|  | Total | Not included | Included | p-value |
| --- | --- | --- | --- | --- |
|  | N=59 | N=21 | N=38 |  |
| Recipient Age | 40 (33-48) | 45 (35-48) | 37 (33-46) | 0.15 |
| Sex |  |  |  | 0.22 |
| Female | 26/59 (44%) | 7/21 (33%) | 19/38 (50%) |  |
| Male | 33/59 (56%) | 14/21 (67%) | 19/38 (50%) |  |
| Race |  |  |  | 0.29 |
| Black African | 55/59 (93%) | 21/21 (100%) | 34/38 (89%) |  |
| Mixed Ancestry | 4/59 (7%) | 0/21 (0%) | 4/38 (11%) |  |

**Table S2:** Comparison between SA kidney donors that are included or not included in study

|  | Total | Not included | Included | p-value |
| --- | --- | --- | --- | --- |
|  | N=32 | N=11 | N=21 |  |
| Donor_age | 30 (26-36) | 30 (24-35) | 31 (26-40) | 0.56 |
| Donor_Sex |  |  |  | 0.68 |
| Female | 9/32 (28%) | 4/11 (36%) | 5/21 (24%) |  |
| Male | 23/32 (72%) | 7/11 (64%) | 16/21 (76%) |  |
| Donor_Race |  |  |  | 0.66 |
| Black African | 19/32 (59%) | 7/11 (64%) | 12/21 (57%) |  |
| Mixed Ancestry | 9/32 (28%) | 2/11 (18%) | 7/21 (33%) |  |
| White | 4/32 (12%) | 2/11 (18%) | 2/21 (10%) |  |

**Table S3:** Comparison between US kidney recipients included or not included in the study

|  | Included  N = 119 | Not Included N = 79 | p-value |
| --- | --- | --- | --- |
| Age, yr, median [IQR] | 55 [46 - 63] | 54 [48 – 61] | 0.9 |
| HIV Positive | 60 (50) | 39 (49) | 1 |
| Race, no. (%): |  |  |  |
| Black or African American | 84 (70) | 60 (76) | 0.2 |
| Other/Mixed ancestry | 10 (8) | 9 (11) |  |
| White | 25 (22) | 10 (13) |  |
| Male sex, no. (%) | 102 (86) | 61 (77) | 0.1 |
| CD4, median [IQR] | 526 [386 - 685] | 478 [366 - 640] | 0.1 |

**Table S4:** Comparison between US kidney donors included or not included in the study

|  | Included  N = 99 | Not Included N = 47 | p-value |
| --- | --- | --- | --- |
| Age, yr, median [IQR] | 36 [28 - 47] | 40 [33 – 49] | 0.06 |
| HIV Positive | 46 (46) | 18 (38) | 0.4 |
| Race, no. (%): |  |  | 0.2 |
| Black or African American | 30 (30) | 14 (30) |  |
| Other/Mixed ancestry | 21 (21) | 5 (10) |  |
| White | 48 (49) | 28 (60) |  |
| Male sex, no. (%) | 69 (70) | 33 (70) | 1 |

**Table S5:** Comparison between kidney recipients from the US HOPE and SA cohorts

|  | US Recipients  N = 119 | SA Recipients N = 43 | p-value |
| --- | --- | --- | --- |
| Age, yr, median [IQR] | 55 [46 - 63] | 37 [33 - 48] | < 0.001 |
| Race, no. (%): |  |  |  |
| Black or African American | 84 (70) | 39 (91) | < 0.001 |
| Other/Mixed ancestry | 10 (8) | 4 (9) |  |
| White | 25 (21) | 0 (0) | < 0.001 |
| Male sex, no. (%) | 102 (85) | 22 (51) |  |
| CD4, median [IQR] | 526 [384 - 686] | 447 [303 – 578] | 0.025 |
| APOL1 RA group, no. (%) |  |  | 0.29 |
| 0 | 44 (37) | 21 (49) |  |
| 1 | 24 (20) | 5 (12) |  |
| 2 | 51 (43) | 17 (39) |  |

**Table S6:** Comparison between kidney donors from the US HOPE and SA cohorts

|  | US Donors  N = 99 | SA Donors N = 21 | p-value |
| --- | --- | --- | --- |
| Age, yr, median [IQR] | 36 [28 - 47] | 31 [26 - 40] | 0.10 |
| HIV Positive | 46 (46) | 21 (100) | - |
| Race, no. (%): |  |  |  |
| Black or African American | 30 (30) | 12 (57) | 0.002 |
| Other/Mixed ancestry | 21 (21) | 7 (33) |  |
| White | 48 (48) | 2 (10) |  |
| Male sex, no. (%) | 69 (69) | 16 (76) | 0.74 |
| CD4, median [IQR] | 406 [284 - 810] | 257 [33 - 805] | 0.11 |
| APOL1 RA group, no. (%) |  |  |  |
| 0 | 50 (88) | 14 (77) | 0.05 |
| 1 | 6 (10) | 7 (33) |  |
| 2 | 1 (1) | 0 (0) |  |

**Table S7:** Comparison between kidney donors from the US HOPE and SA cohorts restricted to HIV+

|  | US Donors  N = 46 | SA Donors N = 21 | p-value |
| --- | --- | --- | --- |
| Age, yr, median [IQR] | 34 [27 - 44] | 31 [26 - 40] | 0.30 |
| Race, no. (%): |  |  | 0.00 |
| Black or African American | 19 (41) | 12 (57) |  |
| Other/Mixed ancestry | 5 (10) | 7 (33) |  |
| White | 22 (47) | 2 (10) |  |
| Male sex, no. (%) | 33 (71) | 16 (76) | 0.9 |
| CD4, median [IQR] | 406 [284 - 810] | 257 [33 - 805] | 0.11 |
| APOL1 RA group, no. (%) |  |  | 0.23 |
| 0 | 33 (82) | 14 (77) |  |
| 1 | 6 (15) | 7 (33) |  |
| 2 | 1 (2) | 0 (0) |  |
