## Supplementary figures and images for "*APOL1* genotype and patient outcomes in US and South African transplant recipients with HIV who received kidneys from donors with HIV"

### Supplemental Figures S1 - S5

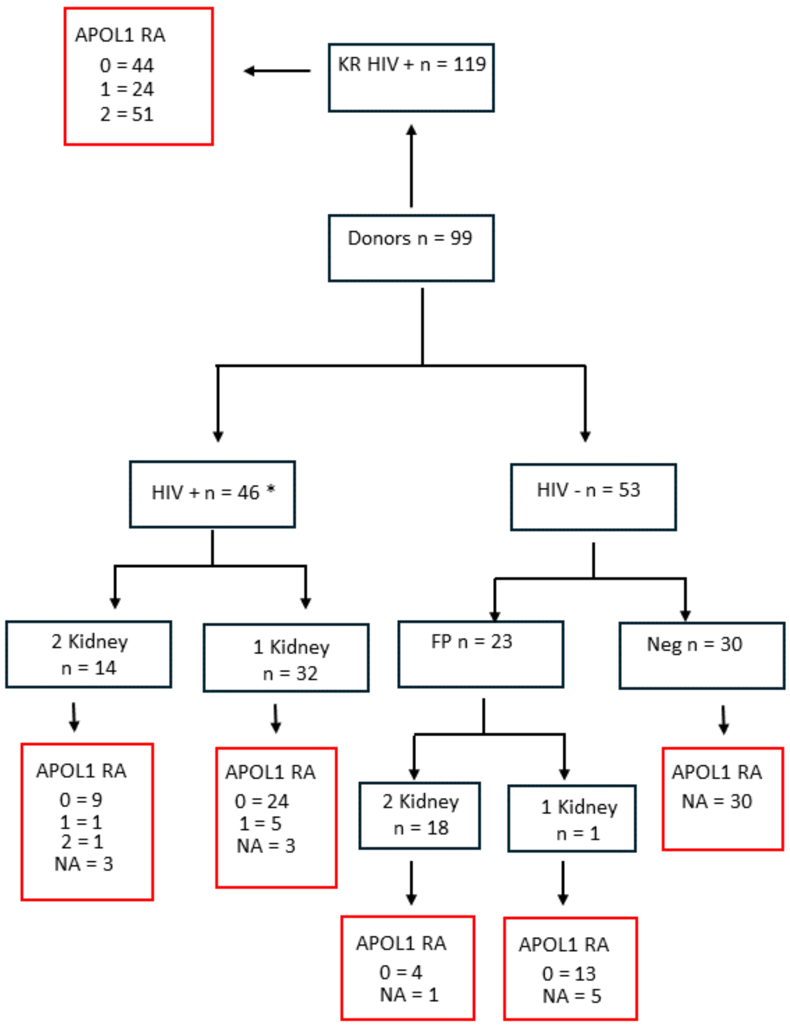

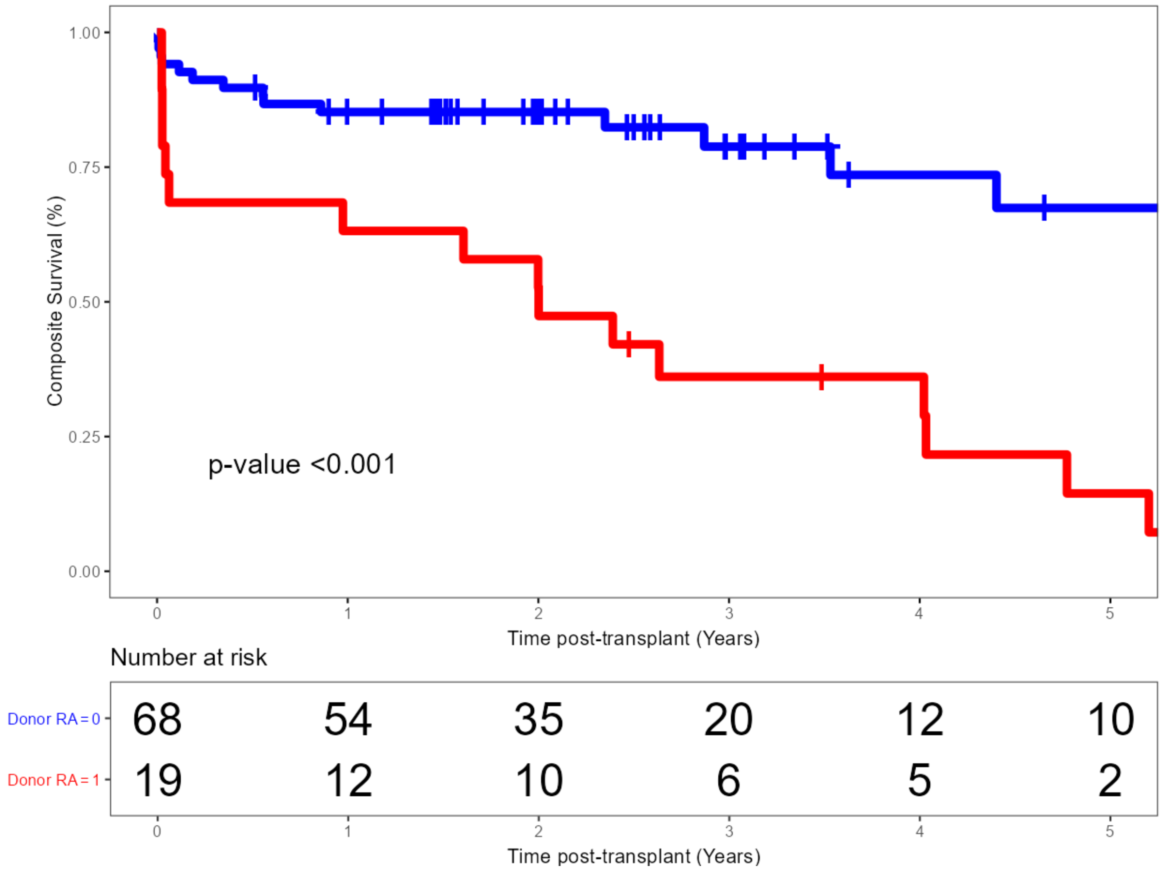

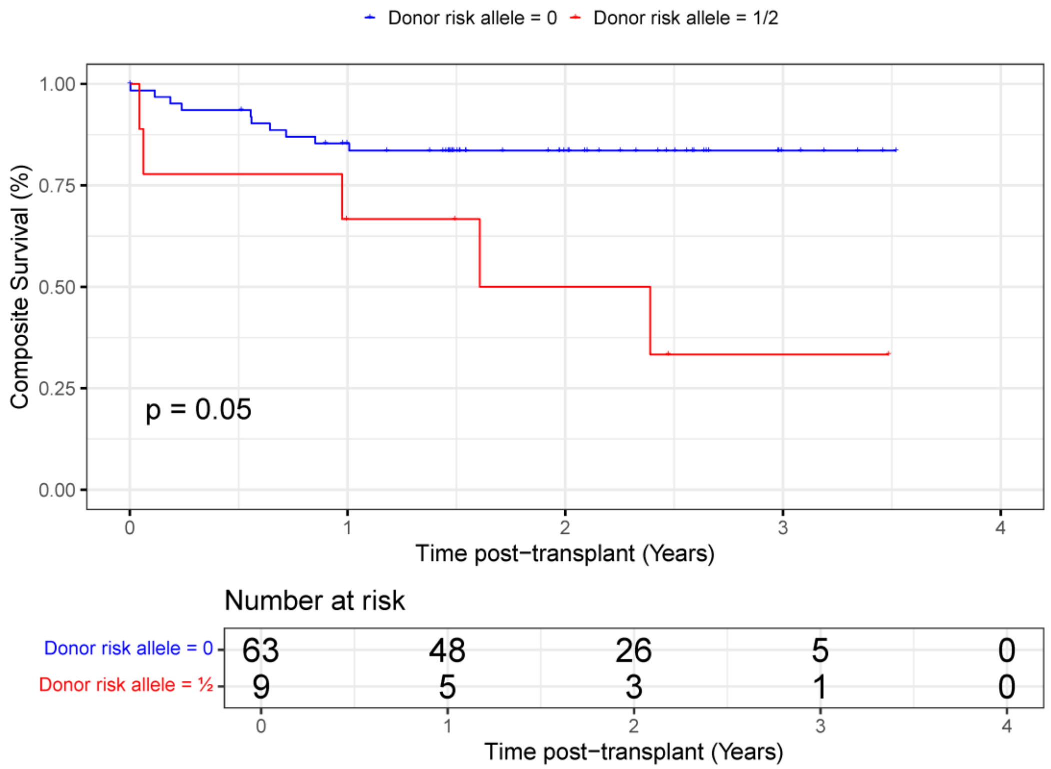

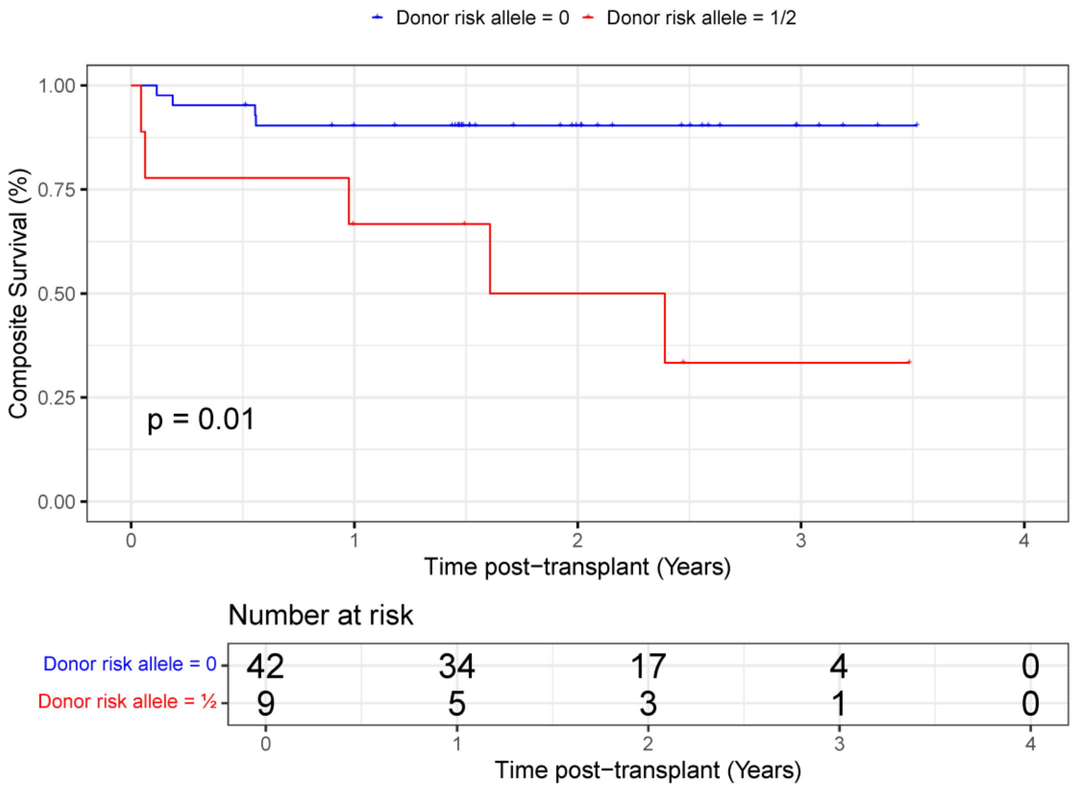

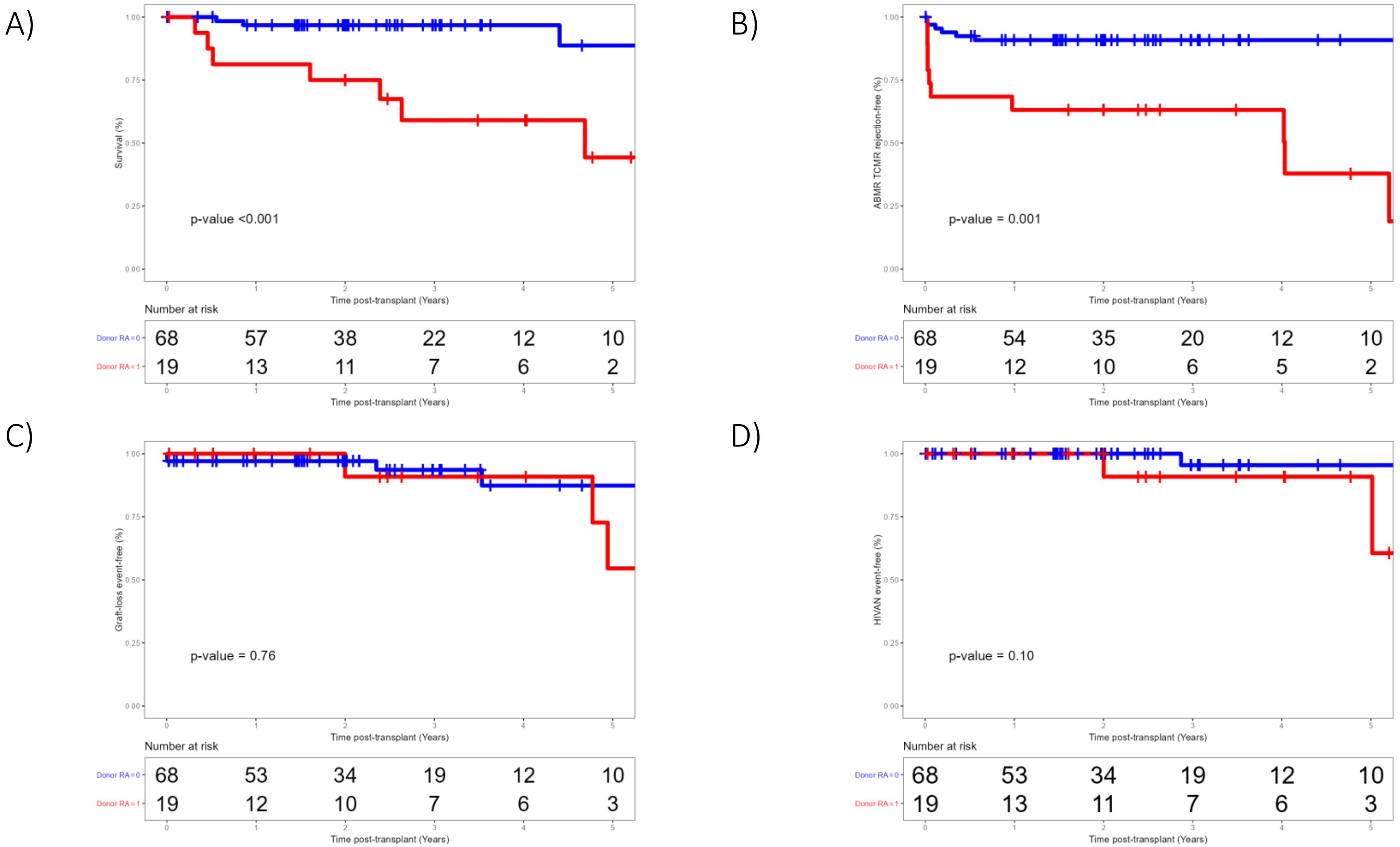
